# Mid-Pregnancy Maternal Leukocyte Telomere Length and Preterm Birth in a Population-Based Hispanic/Latina California Cohort

**DOI:** 10.64898/2026.05.27.26354189

**Authors:** Odessa Garay, Scott Oltman, Rebecca J. Baer, Jue Lin, Janet M. Wojcicki, Kelli K. Ryckman, Laura L. Jelliffe-Pawlowski

## Abstract

**Background:** Preterm birth (PTB) rates among Hispanic/Latina individuals in the United States have risen over the past decade. Data suggest this rise may be driven in part by psychosocial stress. Leukocyte telomere length (LTL), a marker of cumulative cellular aging that shortens under chronic stress, may capture stress-related biological vulnerability, but has not been examined as a potential population-level contributor to PTB in Hispanic/Latina pregnancies.

**Objective:** To examine the association between mid-pregnancy maternal LTL and PTB in a population-based Hispanic/Latina cohort.

**Methods:** In a case-control study nested within a California singleton birth cohort (n = 436 Hispanic/Latina individuals; 215 PTB, 221 term births), LTL was measured by quantitative PCR from biobank specimens collected from 15—20 weeks of gestation. Covariates from linked birth certificate and hospital discharge records were included. Logistic regression estimated ORs and 95% CIs of PTB by LTL examined continuously and by percentile category (≤10th, 11th-89th, ≥90th) with and without adjustment for covariates.

**Results:** Mean and median LTL did not differ between PTB and term births. LTL at or below the 10th percentile was associated with elevated odds of PTB relative to full-term birth (12.6% versus 4.3%; ORc = 3.2, 95% CI 1.3-7.9), persisting after partial (ORadj1 = 3.2, 95% CI 1.3-8.3) and full covariate adjustment (ORadj2 = 3.4, 95% CI 1.3-9.3). Subgroup analyses showed consistent directional patterns across PTB subgroups and for early term birth (ORadj2 = 5.1, 95% CI 1.5-17.0).

**Conclusions:** Mid-pregnancy maternal LTL **≤** 10th percentile was associated with more than three times the odds of PTB, with risk concentrated at the extreme low tail of the distribution. Consistent with a cumulative allostatic load model, markedly short LTL at mid-gestation may reflect elevated stress-related biological risk for preterm delivery. These findings support upstream investment in stress reduction and prospective LTL research in high-burden populations.

## Introduction

Preterm birth (PTB), defined as delivery before 37 completed weeks of gestation, is a leading cause of infant mortality and a major driver of morbidity across the life course [1-3]. In the United States (US), PTB and PTB-related conditions account for more than one-third of all infant deaths [2]. More than one in ten infants in the US is born preterm [4], with substantial variation observed by sociodemographic group including by race/ethnicity and poverty status [4-7].

Hispanic/Latina individuals giving birth, particularly those who are low-income, represent an increasingly vulnerable population with respect to PTB. From 2016 to 2024, the overall PTB rate among Hispanic/Latina births in the US increased from approximately 9.4% to 10.1% (+6.6%), with the largest increase observed among Medicaid-insured individuals born outside the United States, where rates rose from approximately 9.1% to 9.9% (+8.4%) [8]. Over the same period, Hispanic/Latina individuals accounted for a growing share of US births, increasing from approximately 23% in 2016 to more than 27% in 2024, the largest increase observed among major race/ethnicity groups [8]. Together, these trends highlight the increasing contribution of Hispanic/Latina populations to national rates of PTB and underscore the importance of understanding the biological and social pathways that may contribute to risk in this population and to potential in-roads for intervention.

Psychosocial stress, reflected in experiences related to things like adverse life events, discrimination, financial strain, and neighborhood and immigration-related adversity, has been implicated as an important contributor to PTB risk and, in particular, to sociodemographic disparities in birth outcomes [9,10]. Among Hispanic/Latina individuals specifically, immigration-related adversity, including stress among undocumented individuals and sociopolitical stressors linked to anti-immigrant policy environments, has been associated with elevated PTB risk and measurable increases in PTB rates in this population [11-13]. Research suggests that these exposures may influence risk for adverse pregnancy outcomes through processes of biological embedding, whereby repeated or sustained psychosocial stress leads to dysregulation of physiological systems involved in inflammation, oxidative stress, and vascular function [9,10,14].

In research settings, stress is typically operationalized through self-reported measures of perceived stress, anxiety, and depression, through composite indices of cumulative adversity, or through neuroendocrine indicators such as cortisol [9,15]. Leukocyte telomere length (LTL), a measure of the protective DNA caps at the ends of chromosomes, is an established molecular marker of cellular aging and a measurable indicator of cumulative stress burden, as telomere shortening has been linked to oxidative stress and chronic psychosocial adversity [16,17]. Shorter LTL has been linked to stress and stress-related exposures in pregnancy, including factors particularly relevant to Hispanic/Latina populations. In primarily Hispanic/Latina pregnancy cohorts, greater acculturation and acculturative stress during pregnancy have each been associated with shorter LTL [18,19], as have suboptimal gestational weight gain (a correlate of chronic nutritional and psychosocial stress) [20], and maternal hypertension, which also has well-established links to stress-related physiological dysregulation [21].

Beyond its role as a marker of cumulative stress exposure, LTL may also intersect with biological pathways implicated in PTB. For example, shorter LTL has been linked to increased thrombin production and alterations in coagulation pathways [22]. These processes are relevant to PTB because thrombin has been directly implicated in mechanisms leading to preterm premature rupture of membranes (PPROM) [23,24]. Together, these findings suggest that LTL may capture aspects of stress-related biological vulnerability that are relevant to the pathophysiology of PTB.

TL may have potential value not only as a marker of cumulative stress exposure but also as an early indicator of elevated biological risk during pregnancy. Because LTL can be assessed prior to and early in pregnancy it may provide insight into underlying biological susceptibility that precedes PTB-related risks (e.g. gestational hypertension, gestational diabetes) [25,26]. This possibility is particularly relevant for populations experiencing disproportionate and accumulating stress burdens, where earlier identification of heightened biological risk could support targeted monitoring and more timely engagement of supportive care [9,10]. There is also evidence that some stress-related processes related to LTL may be modifiable. Mindfulness-based stress reduction and lifestyle interventions have been associated with preserved or increased LTL in non-pregnant populations [27], and prenatal stress reduction programs have demonstrated reductions in perceived stress, anxiety, and depression during pregnancy [28]. Whether rate of LTL attrition during pregnancy can be altered through such interventions remains an open question, but existing evidence supports the biological plausibility of this pathway [27,29].

Studies examining the association between maternal LTL and PTB have yielded inconsistent findings, with differences in measurement timing, population composition, and sample size likely contributing to the observed heterogeneity [30-32]. Page and colleagues reported that shorter prenatal maternal LTL at 26 to 36 weeks of gestation was associated with PTB in a sample of 100 Mexican-origin women [30]. In contrast, Dutson and colleagues, examining a primarily Latina San Francisco cohort, found longer postpartum maternal LTL associated with moderate to late PTB [31]. A nested case-control study examining postpartum maternal LTL across two diverse cohorts found associations between shorter postpartum LTL and spontaneous PTB [32].

To the best of our knowledge, no prior study has examined mid-pregnancy maternal LTL in relation to PTB in a population-based sample of Hispanic/Latina individuals, assessed whether consideration of demographic and clinical factors alters the observed association, or done so at a gestational window that precedes the onset of the clinical complications that may themselves, be markers of stress-related pathophysiology. This is a critical gap given the potential information provided by examining stress-related LTL shortening in this population and the rising PTB rates among Hispanic/Latina individuals in the US. We address this gap using a population-based cohort of individuals with self-identified Hispanic/Latina ethnicity in California, with LTL measured from banked biospecimens collected at 15 to 20 weeks of gestation as part of routine prenatal screening for aneuploidies, and with access to linked birth certificate and hospital discharge data enabling consideration of a broad range of demographic and clinical factors [33,34].

## Materials and Methods

### Sample

Details regarding the study cohort have been described elsewhere [33-40]. In brief, all participants were drawn from a population-based cohort of singleton births in California from July 2009 through December 2010 (n = 757,853). Gestational dating was confirmed by first-trimester ultrasound. All participants underwent routine first and second trimester serum marker screening for aneuploidies and neural tube defects by the California Genetic Disease Screening Program (n = 241,000). During the second trimester, participants had a serum sample banked by the California Biobank Program (n = 77,604) and had demographic and obstetric information available from birth certificate and hospital discharge records from the California Office of Statewide Health Planning and Development (OSHPD) (n = 61,339). From this cohort, 1,000 banked samples (250 <32 weeks gestational age [GA], 250 32-36 weeks GA, 500 >36 weeks GA) were randomly selected to undergo biomolecular testing. For these analyses, we limited the analytic sample to individuals with self-identified Hispanic/Latina ethnicity (n = 436; Figure 1).

**Figure 1.**
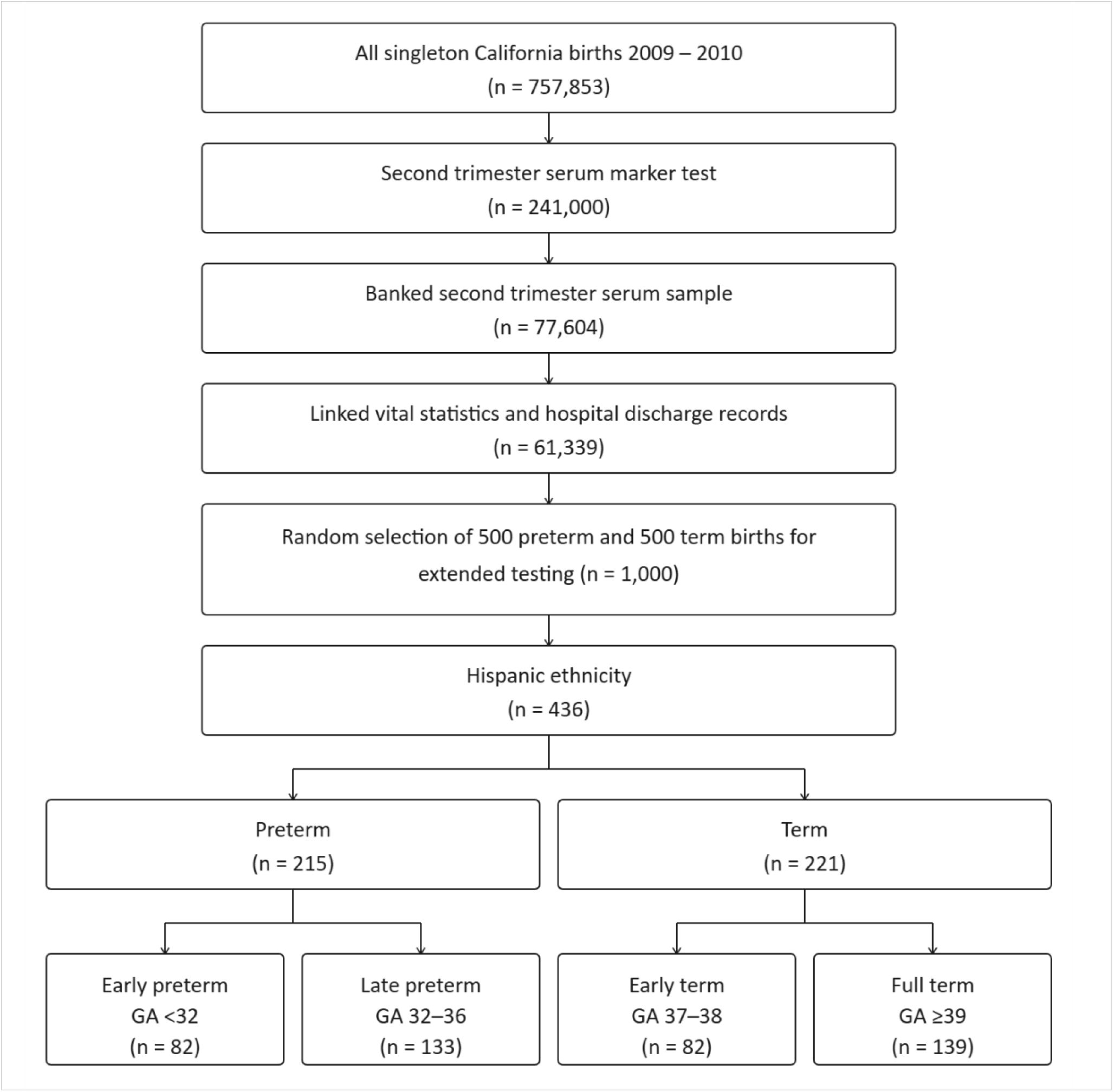
Sample selection.

The primary exposure was second trimester maternal LTL and the outcome of interest was PTB. Covariates included maternal age (<18, 18-34, >34 years), maternal nationality (US-born or non-US-born), Medi-Cal (California’s Medicaid) insurance, participation in the Women, Infants, and Children Supplemental Nutrition Program (WIC), maternal education (<12 years, high school completion, >12 years, or unknown), pre-pregnancy body mass index (BMI) (<18.5, 18.5-24.9, 25-29.9, ≥30), parity (0, 1, ≥2), previous PTB, interpregnancy interval (<6, 6-23, 24-59, ≥60 months), nulliparity (yes versus no), diabetes (preexisting, gestational, or none), hypertension (preexisting, gestational, pre-eclampsia, or none), having a mental health condition, substance abuse (smoking, alcohol, or drug use), and infection. Variables were selected based on availability and prior evidence of association with PTB or LTL [25,41-44]. We note that several adjustment variables, including gestational diabetes, hypertension, BMI, and infection, lie on plausible causal pathways between chronic stress and PTB [45,46]. Their inclusion in fully adjusted models may represent overadjustment, and we address the interpretive implications of this in the Discussion.

### Leukocyte Telomere Length Analysis

DNA was purified from the clotted material using a Quickgene-610L DNA extraction system (Autogen, Holliston, MA, United States). LTL measurement was adapted from Cawthon et al [47]. using quantitative polymerase chain reaction (qPCR). All qPCR was run on the Roche LC480 with 384-tube capacity (Roche Diagnostics Corporation, Indianapolis, IN). The telomere thermal cycling profile (T-PCR) consisted of: Denature at 96°C for 1 minute; denature at 96°C for 1 second, anneal/extend at 54°C for 60 seconds, with fluorescence data collection, 30 cycles. Cycling for single copy gene (S-PCR) consisted of: Denature at 96°C for 1 minute; denature at 95°C for 15 seconds, anneal at 58°C for 1 second, extend at 72°C for 20 seconds, 8 cycles; followed by denature at 96°C for 1 second, anneal at 58°C for 1 second, extend at 72°C for 20 seconds, hold at 83°C for 5 seconds with data collection, 35 cycles. The telomere (T) PCR primer tel1b [5’-CGGTTT(GTTTGG)5GTT-3’] was used at 100 nM concentration and tel2b [5’-GGCTTG(CCTTAC)5CCT-3’] was used at 900 nM concentration. The single-copy gene (S) PCR primers (human β-globin) hbg1 [5’ GCTTCTGACACAACTGTGTTCACTAGC-3’] was used at 300 nM and hbg2 [5’-CACCAACTTCATCCACGTTCACC-3’] was used at 700 nM concentration. The final reaction mix contained 20 mM Tris-HCl, pH 8.4; 50 mM KCl; 200 μM each dNTP; 1% DMSO; 0.4x Syber Green I; 22 ng E. coli DNA; 0.4 Units of Platinum Taq DNA polymerase and ∼ 6 ng of genomic DNA per 11 μl reaction. Tubes containing 26, 8.75, 2.9, 0.97, 0.324 and 0.108ng of a reference DNA and (human genomic DNA, Sigma Aldrich Cat # 11691112001) were included in each PCR run as a standard curve. All samples were measured in triplicate wells in 384 well plates and the average of the T and S values were used to calculate the T/S ratios after removal of outliners with the Dixon Q test. The T/S ratio for each sample was measured twice, each in triplicate wells. If the duplicate T/S value and initial value varied by more than 7%, the sample was run a third time, and the two closest values was be reported. 20.7% of samples were run a third time. The CV for this study was 2.2±1.5%. Further protocol details are available from the Telomere Research Network (https://trn.tulane.edu/resources/lab-protocols/).

### Statistical Analyses

Demographic and clinical characteristics were compared between pregnancies with PTB and full-term birth using t-tests and chi-squared tests for continuous and categorical variables, respectively. Maternal LTL was compared across outcome groups using within-group means and medians and by percentile category (≤10th, 11th-89th, ≥90th percentile), with cut points determined using all subjects. Odds ratios (ORs) and 95% confidence intervals (CIs) for LTL-outcome associations were estimated using logistic regression. Crude ORs (ORc) were generated for univariable assessments. Adjusted ORs (ORadj) were generated in two stages: first including only demographic and clinical factors found to be associated with PTB (ORadj1), then including all candidate variables (ORadj2). Exploratory analyses compared LTL across early PTB, late PTB, and early term birth relative to full-term birth. All analyses were performed using SAS 9.4 (SAS Institute, Cary, NC).

### Ethics and Data Availability

Methods and protocols were approved by the Committee for the Protection of Human Subjects within the California Health and Human Services Agency and the Institutional Review Board of the University of California, San Francisco. Use of the California Biobank Program specimens and California birth certificate and OSHPD hospital discharge data was conducted under data use agreements with the California Department of Public Health. Individual written informed consent was not required for this secondary analysis of de-identified administrative and biobank data. California law requires that all investigators using individual-level data obtain approval for use. As such, while these data are not opening available for use by other investigators, individuals can apply for data use and biospecimens from the California Department of Public Health.

## Results

The analytic sample included 436 individuals: 82 with early PTB (<32 weeks), 133 with moderate/late PTB (32-36 weeks), 82 with early term birth (37-38 weeks), and 139 with full-term birth (39+ weeks) (Table 1, Table 2). Across all groups, participants were predominantly between 18 and 34 years of age, enrolled in Medi-Cal, and approximately half were foreign-born. WIC participation was similarly high across groups (65.9-74.4%) (Table 1). Sociodemographic characteristics including maternal age, nationality, insurance status, education, parity, and interpregnancy interval did not differ significantly between any gestational age group and the full-term reference group (Table 2). Mean pre-pregnancy BMI was similar across all groups (26.3-27.4). Pregnancies ending in PTB differed from full-term births primarily in the frequency of gestational diabetes (18.1% versus 7.2%; ORc = 3.0, 95% CI 1.4-6.2), pre-eclampsia (18.6% versus 2.9%; ORc = 8.0, 95% CI 2.8-23.0), and infection (9.3% versus 2.9%; ORc = 3.5, 95% CI 1.2-10.4) (Table 2). The clinical profile differed meaningfully by subgroup. Early PTB carried the broadest burden: pre-eclampsia (25.6% versus 2.9%; ORc = 12.3, 95% CI 4.0-37.4) and infection (19.5% versus 2.9%; ORc = 8.2, 95% CI 2.6-25.4) showed the strongest associations, and preexisting diabetes (6.1% versus 0.7%; ORc = 10.3, 95% CI 1.2-90.3) and low BMI (<18.5: 11.0% versus 4.3%; ORc = 3.8, 95% CI 1.2-11.9) were significantly elevated, findings not observed in other subgroups. Late PTB was characterized by elevated gestational diabetes (18.1% versus 7.2%; ORc = 2.9, 95% CI 1.3-6.4) and pre-eclampsia (14.3% versus 2.9%; ORc = 5.8, 95% CI 1.9-17.6), but infection was not elevated (3.0% versus 2.9%). Early term birth did not differ significantly from full-term birth on any clinical condition examined.

**Table 1.**
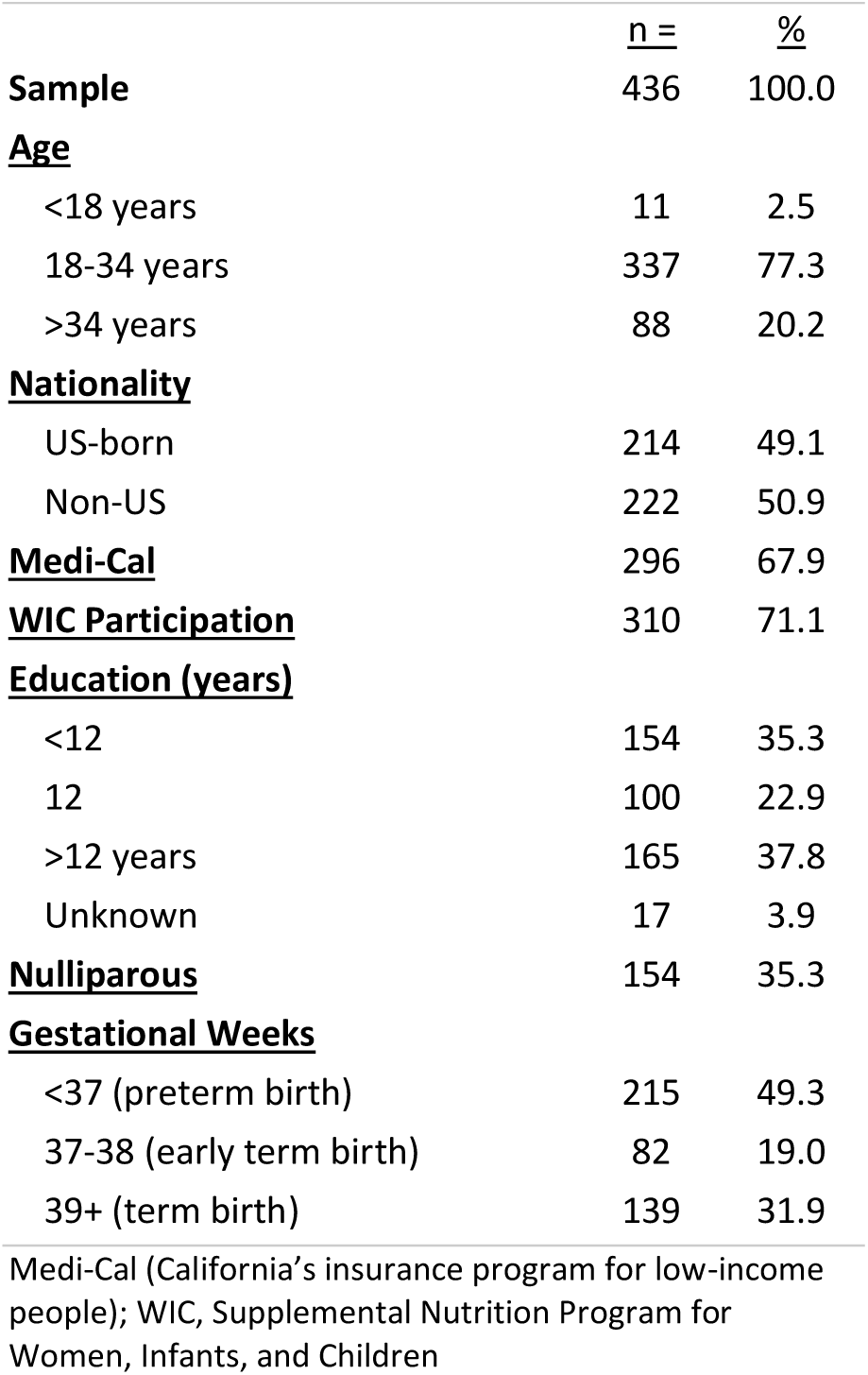
Sample overview.

|  | <u>n =</u> | <u>%</u> |
| --- | --- | --- |
| **Sample** | 436 | 100.0 |
| **<u>Age</u>** |  |  |
| <18 years | 11 | 2.5 |
| 18-34 years | 337 | 77.3 |
| >34 years | 88 | 20.2 |
| **<u>Nationality</u>** |  |  |
| US-born | 214 | 49.1 |
| Non-US | 222 | 50.9 |
| **<u>Medi-Cal</u>** | 296 | 67.9 |
| **<u>WIC Participation</u>** | 310 | 71.1 |
| **<u>Education (years)</u>** |  |  |
| <12 | 154 | 35.3 |
| 12 | 100 | 22.9 |
| >12 years | 165 | 37.8 |
| Unknown | 17 | 3.9 |
| **<u>Nulliparous</u>** | 154 | 35.3 |
| **<u>Gestational Weeks</u>** |  |  |
| <37 (preterm birth) | 215 | 49.3 |
| 37-38 (early term birth) | 82 | 19.0 |
| 39+ (term birth) | 139 | 31.9 |

Medi-Cal (California's insurance program for low-income people); WIC, Supplemental Nutrition Program for Women, Infants, and Children

**Table 2.**
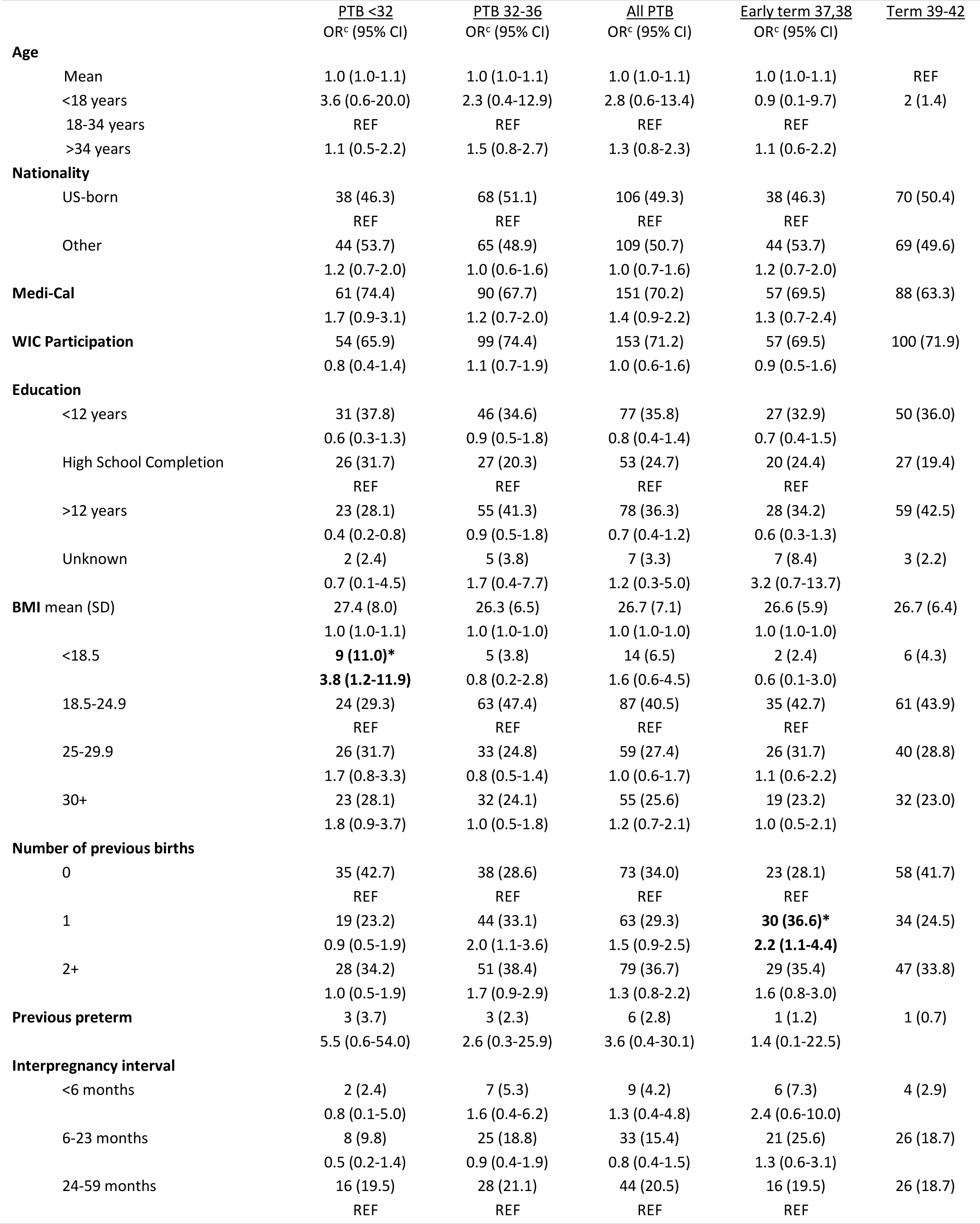
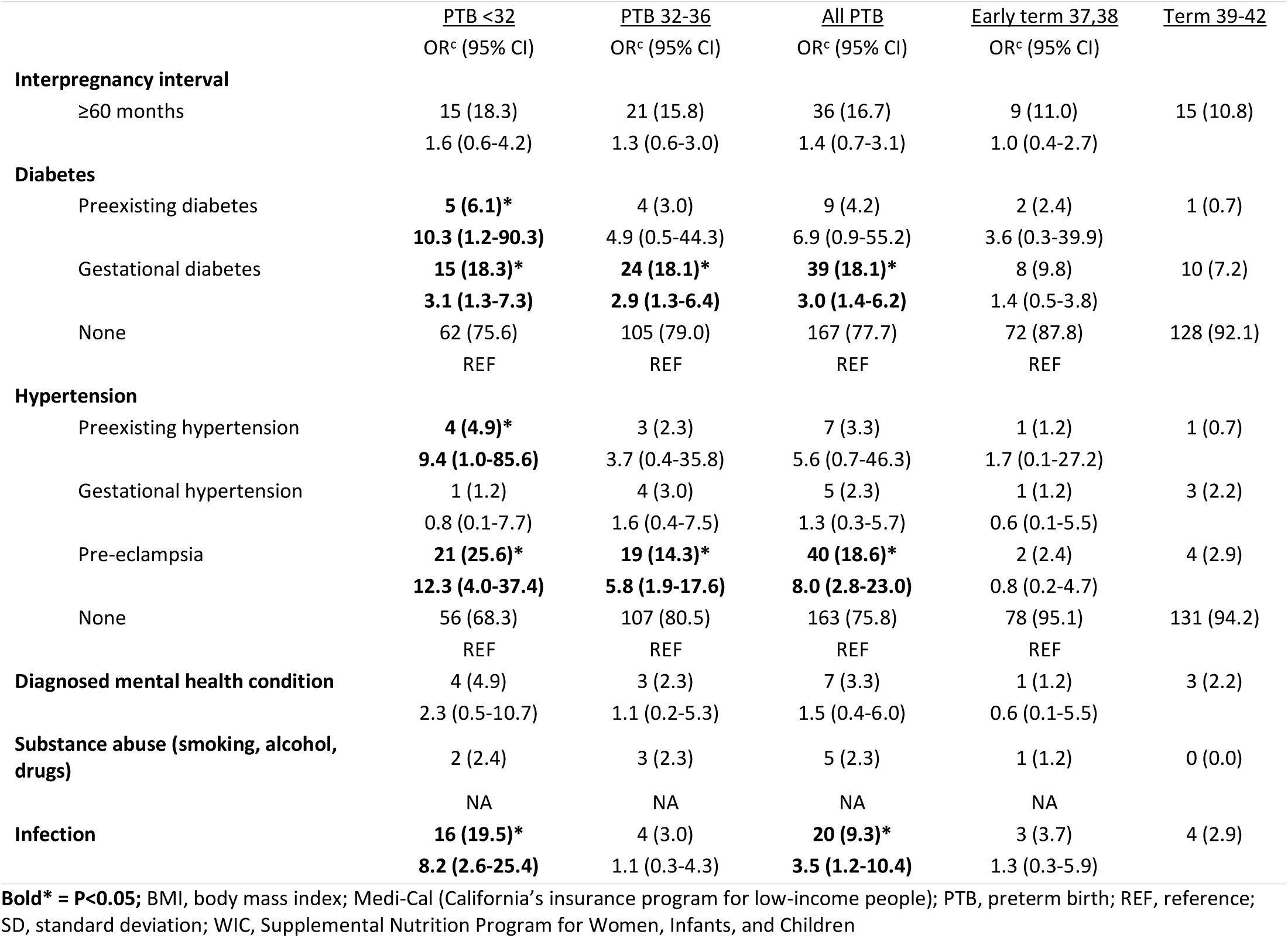
Association of maternal variables with gestational age categories.

|  | <u>PTB <32</u><br>OR<sup>c</sup> (95% CI) | <u>PTB 32-36</u><br>OR<sup>c</sup> (95% CI) | <u>All PTB</u><br>OR<sup>c</sup> (95% CI) | <u>Early term 37,38</u><br>OR<sup>c</sup> (95% CI) | <u>Term 39-42</u> |
| --- | --- | --- | --- | --- | --- |
| **Age** |  |  |  |  |  |
| Mean | 1.0 (1.0-1.1) | 1.0 (1.0-1.1) | 1.0 (1.0-1.1) | 1.0 (1.0-1.1) | REF |
| <18 years | 3.6 (0.6-20.0) | 2.3 (0.4-12.9) | 2.8 (0.6-13.4) | 0.9 (0.1-9.7) | 2 (1.4) |
| 18-34 years | REF | REF | REF | REF |  |
| >34 years | 1.1 (0.5-2.2) | 1.5 (0.8-2.7) | 1.3 (0.8-2.3) | 1.1 (0.6-2.2) |  |
| **Nationality** |  |  |  |  |  |
| US-born | 38 (46.3) | 68 (51.1) | 106 (49.3) | 38 (46.3) | 70 (50.4) |
|  | REF | REF | REF | REF |  |
| Other | 44 (53.7) | 65 (48.9) | 109 (50.7) | 44 (53.7) | 69 (49.6) |
|  | 1.2 (0.7-2.0) | 1.0 (0.6-1.6) | 1.0 (0.7-1.6) | 1.2 (0.7-2.0) |  |
| **Medi-Cal** | 61 (74.4) | 90 (67.7) | 151 (70.2) | 57 (69.5) | 88 (63.3) |
|  | 1.7 (0.9-3.1) | 1.2 (0.7-2.0) | 1.4 (0.9-2.2) | 1.3 (0.7-2.4) |  |
| **WIC Participation** | 54 (65.9) | 99 (74.4) | 153 (71.2) | 57 (69.5) | 100 (71.9) |
|  | 0.8 (0.4-1.4) | 1.1 (0.7-1.9) | 1.0 (0.6-1.6) | 0.9 (0.5-1.6) |  |
| **Education** |  |  |  |  |  |
| <12 years | 31 (37.8) | 46 (34.6) | 77 (35.8) | 27 (32.9) | 50 (36.0) |
|  | 0.6 (0.3-1.3) | 0.9 (0.5-1.8) | 0.8 (0.4-1.4) | 0.7 (0.4-1.5) |  |
| High School Completion | 26 (31.7) | 27 (20.3) | 53 (24.7) | 20 (24.4) | 27 (19.4) |
|  | REF | REF | REF | REF |  |
| >12 years | 23 (28.1) | 55 (41.3) | 78 (36.3) | 28 (34.2) | 59 (42.5) |
|  | 0.4 (0.2-0.8) | 0.9 (0.5-1.8) | 0.7 (0.4-1.2) | 0.6 (0.3-1.3) |  |
| Unknown | 2 (2.4) | 5 (3.8) | 7 (3.3) | 7 (8.4) | 3 (2.2) |
|  | 0.7 (0.1-4.5) | 1.7 (0.4-7.7) | 1.2 (0.3-5.0) | 3.2 (0.7-13.7) |  |
| **BMI mean (SD)** | 27.4 (8.0) | 26.3 (6.5) | 26.7 (7.1) | 26.6 (5.9) | 26.7 (6.4) |
|  | 1.0 (1.0-1.1) | 1.0 (1.0-1.0) | 1.0 (1.0-1.0) | 1.0 (1.0-1.0) |  |
| <18.5 | **9 (11.0)\*** | 5 (3.8) | 14 (6.5) | 2 (2.4) | 6 (4.3) |
|  | **3.8 (1.2-11.9)** | 0.8 (0.2-2.8) | 1.6 (0.6-4.5) | 0.6 (0.1-3.0) |  |
| 18.5-24.9 | 24 (29.3) | 63 (47.4) | 87 (40.5) | 35 (42.7) | 61 (43.9) |
|  | REF | REF | REF | REF |  |
| 25-29.9 | 26 (31.7) | 33 (24.8) | 59 (27.4) | 26 (31.7) | 40 (28.8) |
|  | 1.7 (0.8-3.3) | 0.8 (0.5-1.4) | 1.0 (0.6-1.7) | 1.1 (0.6-2.2) |  |
| 30+ | 23 (28.1) | 32 (24.1) | 55 (25.6) | 19 (23.2) | 32 (23.0) |
|  | 1.8 (0.9-3.7) | 1.0 (0.5-1.8) | 1.2 (0.7-2.1) | 1.0 (0.5-2.1) |  |
| **Number of previous births** |  |  |  |  |  |
| 0 | 35 (42.7) | 38 (28.6) | 73 (34.0) | 23 (28.1) | 58 (41.7) |
|  | REF | REF | REF | REF |  |
| 1 | 19 (23.2) | 44 (33.1) | 63 (29.3) | **30 (36.6)\*** | 34 (24.5) |
|  | 0.9 (0.5-1.9) | 2.0 (1.1-3.6) | 1.5 (0.9-2.5) | **2.2 (1.1-4.4)** |  |
| 2+ | 28 (34.2) | 51 (38.4) | 79 (36.7) | 29 (35.4) | 47 (33.8) |
|  | 1.0 (0.5-1.9) | 1.7 (0.9-2.9) | 1.3 (0.8-2.2) | 1.6 (0.8-3.0) |  |
| **Previous preterm** | 3 (3.7) | 3 (2.3) | 6 (2.8) | 1 (1.2) | 1 (0.7) |
|  | 5.5 (0.6-54.0) | 2.6 (0.3-25.9) | 3.6 (0.4-30.1) | 1.4 (0.1-22.5) |  |
| **Interpregnancy interval** |  |  |  |  |  |
| <6 months | 2 (2.4) | 7 (5.3) | 9 (4.2) | 6 (7.3) | 4 (2.9) |
|  | 0.8 (0.1-5.0) | 1.6 (0.4-6.2) | 1.3 (0.4-4.8) | 2.4 (0.6-10.0) |  |
| 6-23 months | 8 (9.8) | 25 (18.8) | 33 (15.4) | 21 (25.6) | 26 (18.7) |
|  | 0.5 (0.2-1.4) | 0.9 (0.4-1.9) | 0.8 (0.4-1.5) | 1.3 (0.6-3.1) |  |
| 24-59 months | 16 (19.5) | 28 (21.1) | 44 (20.5) | 16 (19.5) | 26 (18.7) |
|  | REF | REF | REF | REF |  |

**Table 2** (continued)
|  | <u>PTB <32</u> | <u>PTB 32-36</u> | <u>All PTB</u> | <u>Early term 37,38</u> | <u>Term 39-42</u> |
| --- | --- | --- | --- | --- | --- |
|  | OR<sup>c</sup> (95% CI) | OR<sup>c</sup> (95% CI) | OR<sup>c</sup> (95% CI) | OR<sup>c</sup> (95% CI) |  |
| **Interpregnancy interval** |  |  |  |  |  |
| ≥60 months | 15 (18.3) | 21 (15.8) | 36 (16.7) | 9 (11.0) | 15 (10.8) |
|  | 1.6 (0.6-4.2) | 1.3 (0.6-3.0) | 1.4 (0.7-3.1) | 1.0 (0.4-2.7) |  |
| **Diabetes** |  |  |  |  |  |
| Preexisting diabetes | **5 (6.1)\*** | 4 (3.0) | 9 (4.2) | 2 (2.4) | 1 (0.7) |
|  | **10.3 (1.2-90.3)** | 4.9 (0.5-44.3) | 6.9 (0.9-55.2) | 3.6 (0.3-39.9) |  |
| Gestational diabetes | **15 (18.3)\*** | **24 (18.1)\*** | **39 (18.1)\*** | 8 (9.8) | 10 (7.2) |
|  | **3.1 (1.3-7.3)** | **2.9 (1.3-6.4)** | **3.0 (1.4-6.2)** | 1.4 (0.5-3.8) |  |
| None | 62 (75.6) | 105 (79.0) | 167 (77.7) | 72 (87.8) | 128 (92.1) |
|  | REF | REF | REF | REF |  |
| **Hypertension** |  |  |  |  |  |
| Preexisting hypertension | **4 (4.9)\*** | 3 (2.3) | 7 (3.3) | 1 (1.2) | 1 (0.7) |
|  | **9.4 (1.0-85.6)** | 3.7 (0.4-35.8) | 5.6 (0.7-46.3) | 1.7 (0.1-27.2) |  |
| Gestational hypertension | 1 (1.2) | 4 (3.0) | 5 (2.3) | 1 (1.2) | 3 (2.2) |
|  | 0.8 (0.1-7.7) | 1.6 (0.4-7.5) | 1.3 (0.3-5.7) | 0.6 (0.1-5.5) |  |
| Pre-eclampsia | **21 (25.6)\*** | **19 (14.3)\*** | **40 (18.6)\*** | 2 (2.4) | 4 (2.9) |
|  | **12.3 (4.0-37.4)** | **5.8 (1.9-17.6)** | **8.0 (2.8-23.0)** | 0.8 (0.2-4.7) |  |
| None | 56 (68.3) | 107 (80.5) | 163 (75.8) | 78 (95.1) | 131 (94.2) |
|  | REF | REF | REF | REF |  |
| **Diagnosed mental health condition** | 4 (4.9) | 3 (2.3) | 7 (3.3) | 1 (1.2) | 3 (2.2) |
|  | 2.3 (0.5-10.7) | 1.1 (0.2-5.3) | 1.5 (0.4-6.0) | 0.6 (0.1-5.5) |  |
| **Substance abuse (smoking, alcohol, drugs)** | 2 (2.4) | 3 (2.3) | 5 (2.3) | 1 (1.2) | 0 (0.0) |
|  | NA | NA | NA | NA |  |
| **Infection** | **16 (19.5)\*** | 4 (3.0) | **20 (9.3)\*** | 3 (3.7) | 4 (2.9) |
|  | **8.2 (2.6-25.4)** | 1.1 (0.3-4.3) | **3.5 (1.2-10.4)** | 1.3 (0.3-5.9) |  |

**Bold\*** = **P<0.05**; BMI, body mass index; Medi-Cal (California's insurance program for low-income people); PTB, preterm birth; REF, reference; SD, standard deviation; WIC, Supplemental Nutrition Program for Women, Infants, and Children

When examined as a continuous variable, LTL was found to be similar across all outcome groups (Table 3, Figure 2). When LTL was categorized by percentile, individuals with PTB were more likely than those with full-term birth to have a LTL at or below the 10th percentile (12.6% versus 4.3%; ORc = 3.2, 95% CI 1.3-7.9). This association persisted after partial adjustment (OR adj1 = 3.2, 95% CI 1.3-8.3) and strengthened with full covariate adjustment (OR adj2 = 3.4, 95% CI 1.3-9.3). LTL at or above the 90th percentile was not significantly associated with PTB in any model. In subgroup analyses, the association between LTL at or below the 10th percentile and outcome was directionally consistent across gestational age groups. For late PTB the association was statistically significant (crude OR = 3.4, 95% CI 1.3-9.0; OR adj2 = 5.0, 95% CI 1.7-14.9), as was the association with early term birth (crude OR = 3.0, 95% CI 1.03-8.5; OR adj2 = 5.1, 95% CI 1.5-17.0). For early PTB the direction was consistent but the estimate was imprecise and confidence intervals crossed 1.0 in all models (OR crude = 2.7, 95% CI 0.9-7.9; ORadj2 = 3.3, 95% CI 0.8-13.2).

**Figure 2.**
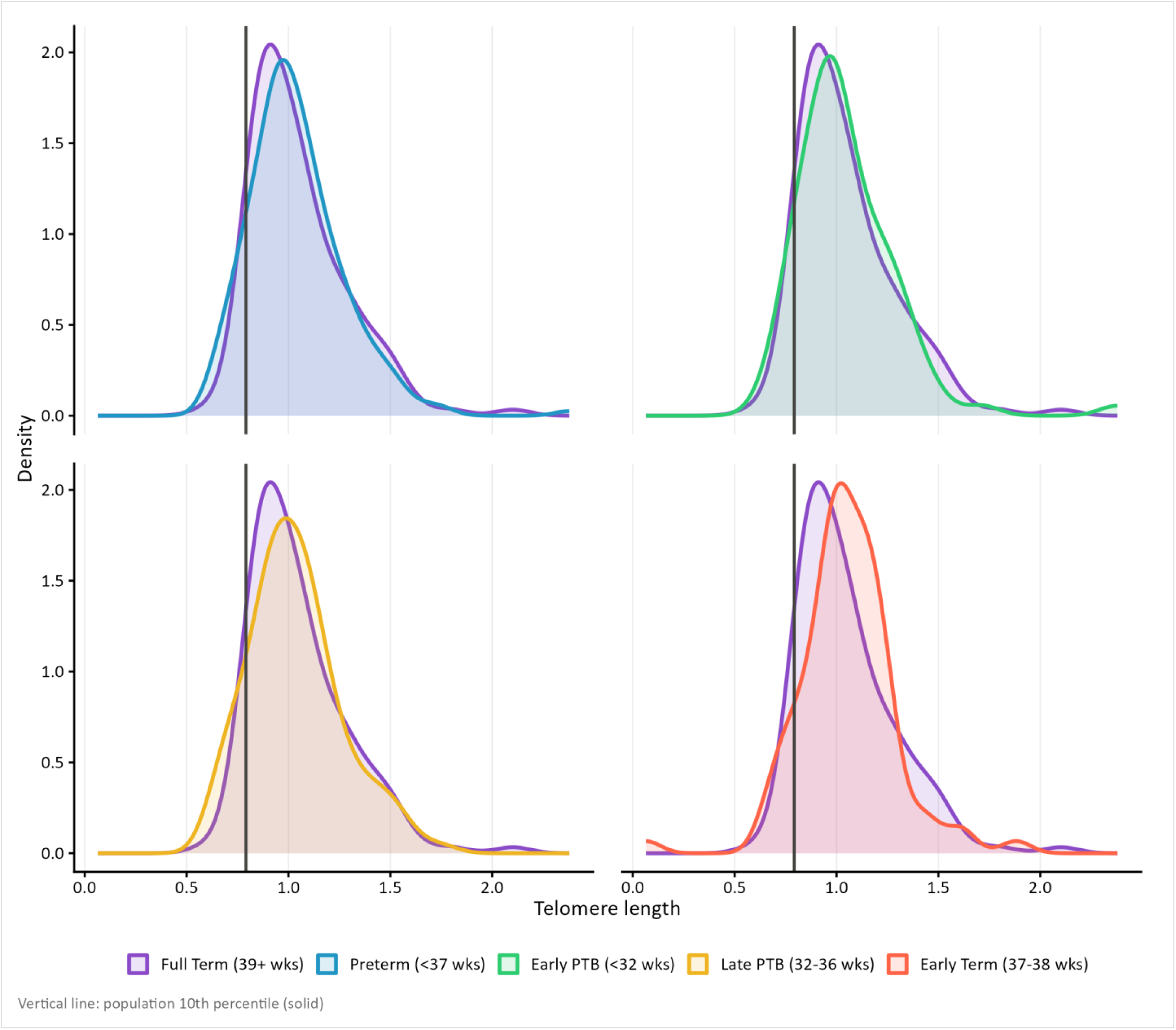
Distribution density plots of second trimester leukocyte telomere length by gestational age group. Panels display: all preterm birth (PTB, <37 weeks; upper left), early PTB (<32 weeks; upper right), late PTB (32-36 weeks; bottom left), and early term births (37-38 weeks; bottom right) vs full term births (39+ weeks). The vertical line represents the population 10^th^ percentile of leukocyte telomere length.

**Figure 3.**
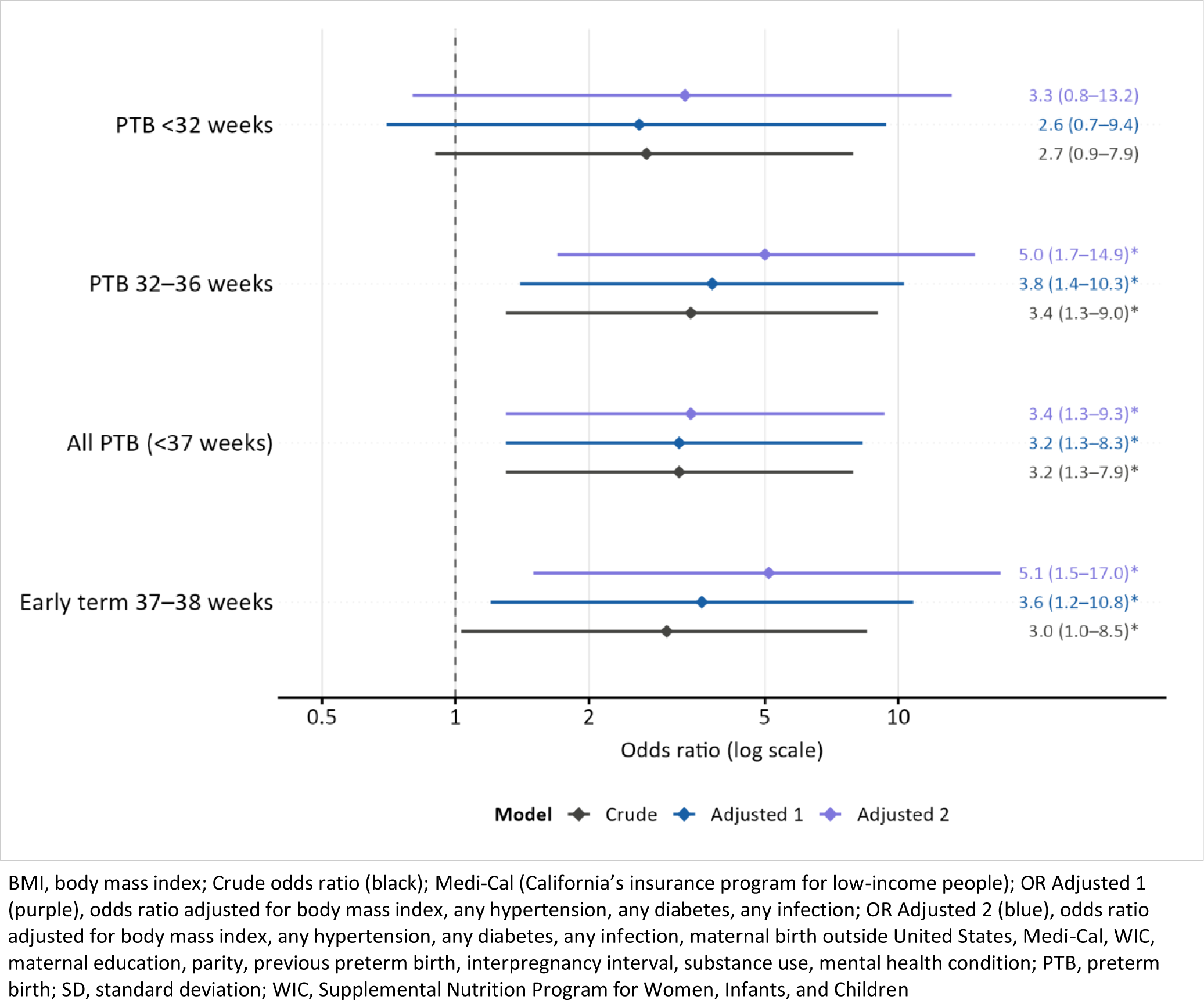
Forest plot of odds ratios with 95% confidence intervals for models of gestational age category by leukocyte telomere length.

**Table 3.**
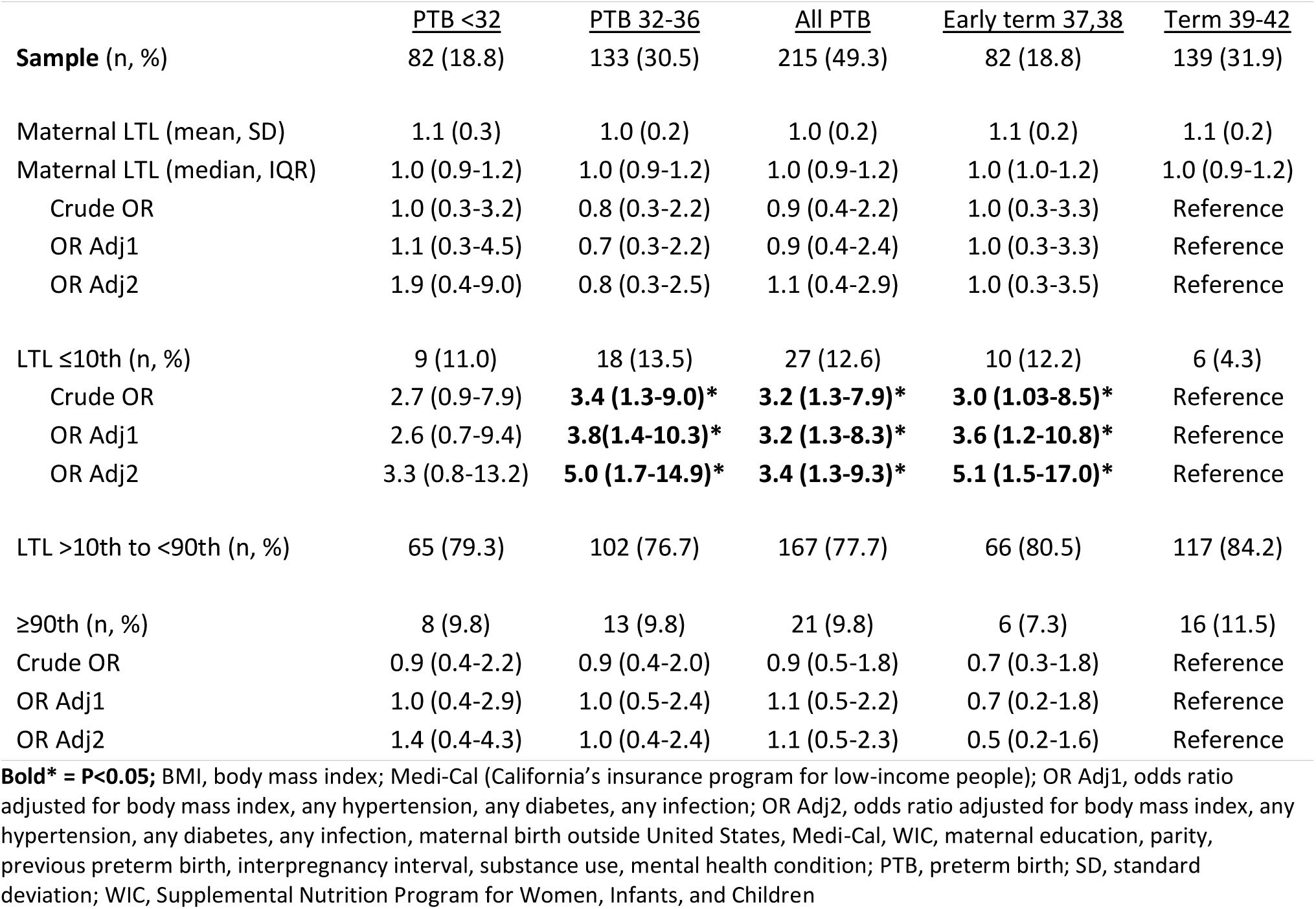
Association of leukocyte telomere length (LTL) and preterm birth by group.

## Discussion

In this population-based cohort of Hispanic/Latina individuals in California, we found that mid-pregnancy maternal LTL at or below the 10th percentile was associated with more than three times the odds of PTB, and that this association persisted and strengthened with covariate adjustment. The signal was concentrated at the extreme low tail of the LTL distribution: mean and median LTL did not differ across outcome groups, indicating that what distinguishes those who delivered preterm is not likely a modest generalized shortening but rather, perhaps, the presence of markedly short LTL at mid-gestation. This distribution pattern is consistent with a threshold or cumulative burden model, in which biological vulnerability becomes detectable primarily among those carrying the heaviest allostatic load [48,49]. A similar directional pattern was observed for late PTB and for early term birth, though the early PTB subgroup was too small for reliable estimation. To our knowledge, this is the largest population-based study to date examining this association in a Hispanic/Latina cohort, and the first to assess whether covariate adjustment alters the observed relationship.

### Results in Context of What is Known

The pattern we observed, with PTB risk concentrated at the extreme low end of the LTL distribution, has a plausible biological basis rooted in the cumulative nature of stress-related cellular aging. The allostatic load framework proposes that repeated exposure to psychosocial and structural adversity leads to accelerating wear on physiological systems, with biological aging markers including LTL reflecting this accumulation over time [48,49,50]. In Hispanic/Latina populations, this process is described as weathering, whereby life-course exposure to structural disadvantage, discrimination, and poverty accelerates biological aging, has been invoked to explain the observed divergence between foreign-born and US-born Latina women’s birth outcomes [51,52]. Our finding that the LTL-PTB association is concentrated in the lowest decile is consistent with this framework: individuals at the 10th percentile of LTL may represent those for whom cumulative stress exposures have crossed a biological threshold that affects gestational physiology, while those with moderately shortened LTL have not yet reached that threshold [48,49].

Our findings align in direction with those of Page and colleagues, who reported shorter prenatal maternal LTL associated with PTB in 100 Mexican-origin women, though they observed a shift in median LTL while we did not [30]. This difference may reflect the larger, more heterogeneous stress exposure profile of our population-based sample compared with a recruited clinical cohort, or may suggest that the distributional pattern of LTL shortening, rather than central tendency, is the more relevant signal. A pilot study in African American women similarly found shorter maternal peripheral blood LTL associated with PTB, though that study was conducted later in gestation [53]. Dutson and colleagues found longer postpartum maternal LTL associated with PTB in a primarily Latina San Francisco cohort [31].

The divergence in direction between prenatal studies, which generally find shorter LTL associated with PTB, and postpartum studies, which have found the opposite, may reflect the different biological processes indexed at each measurement window. A pilot longitudinal study of LTL dynamics across uncomplicated pregnancies found no significant within-person change in LTL across gestation, but did find shorter postpartum LTL among those who delivered by cesarean section [54], suggesting that mode of delivery and peripartum inflammatory processes may introduce systematic measurement artifacts in postpartum samples. Panelli and colleagues found associations between shorter postpartum LTL and spontaneous PTB across two diverse cohorts, further illustrating the complexity of measurement timing in this literature [32].

The broader literature on LTL as a biomarker of stress and adversity provides additional context for interpreting our findings. LTL has been consistently associated with socioeconomic disadvantage, discrimination, and chronic psychosocial stress across diverse populations [49,55,56], and LTL shortening accelerates under conditions of high allostatic load [49,50]. In our data, gestational diabetes, pre-eclampsia, and infection were each significantly more common among those with PTB, and each of these conditions has established biological links to oxidative stress and inflammation, the same pathways that drive LTL shortening [9,10,14]. The co-occurrence of short LTL with conditions reflecting systemic oxidative and inflammatory burden is consistent with a shared upstream pathway, in which accumulated stress exposure manifests simultaneously in LTL attrition and in the clinical conditions that independently elevate PTB risk.

### Pathophysiological Mechanisms

Two mechanistic pathways may underlie the association between short mid-pregnancy maternal LTL and PTB, and they are not mutually exclusive. The first operates through the stress-oxidative damage axis. LTL shortens under conditions of cumulative oxidative stress and chronic inflammation, and these same processes dysregulate the physiological systems governing gestational maintenance [16,17]. A compelling biological framework for linking LTL directly to the timing of parturition has been proposed by Phillippe, who describes a gestational telomere clock in which LTL in placental and fetal membrane tissues shortens progressively throughout gestation, and in which conditions that accelerate oxidative stress, including maternal psychosocial stress, bring LTL to a critically short threshold earlier than expected, triggering the cellular senescence cascades that initiate preterm labor [57]. Under this model, shorter maternal leukocyte LTL at mid-gestation may serve as a systemic marker of an oxidative stress environment that is simultaneously shortening LTL in gestational tissues and advancing the parturition clock [57,58]. The second pathway involves coagulation: shorter LTL has been associated with increased thrombin production [22,23], and thrombin promotes fetal membrane weakening through matrix metalloproteinase activation and inflammatory signaling, with particularly strong links to PPROM [24].

Direct evidence supporting the LTL-PPROM link comes from studies finding shorter fetal leukocyte LTL in pregnancies complicated by PPROM compared with PTB with intact membranes [59]. These two pathways converge on a common theme: LTL shortening as a marker of a systemic oxidative and inflammatory environment that, when sufficiently advanced at mid-pregnancy, manifests in both placental-fetal membrane dysfunction and elevated PTB risk. The strengthening of the association with full covariate adjustment in our data is consistent with this framework but warrants caution: several adjustment variables lie on causal pathways between stress and PTB [45,46], and the strengthening may reflect negative confounding rather than unmixed pathophysiology.

### Clinical and Public Health Implications

These findings have several implications for clinical care and public health practice in Hispanic/Latina populations, where PTB rates have risen substantially alongside documented increases in stress-related risk factors [8]. LTL is not currently a validated clinical risk marker: no established threshold exists for clinical use, and the qPCR-based assay used here requires banked research-grade specimens rather than point-of-care measurement [47]. However, the concentration of PTB risk at the 10th percentile of LTL, measured at 15 to 20 weeks before gestational complications have typically emerged, suggests that if LTL measurement were validated prospectively in diverse populations, it could offer a biologically grounded signal for stratifying monitoring intensity earlier in pregnancy [25,26]. For clinical care at present, the more proximate implications lie in the clinical findings themselves. The strong associations of pre-eclampsia and gestational diabetes with PTB in our data underscore the established importance of early prenatal care for timely identification and management of these conditions [60]. Among Hispanic/Latina individuals, documented barriers to prenatal care including language barriers, discrimination within healthcare settings, and immigration-related fear of healthcare contact [61,62] represent structural drivers of delayed or inadequate care that are more immediately actionable than any biomarker-based strategy.

At the upstream level, the allostatic load framework suggests that interventions aimed at reducing the cumulative stress burden carried into pregnancy may have the greatest potential for population-level impact [48,49]. Prenatal stress reduction programs have demonstrated reductions in perceived stress, anxiety, and depression during pregnancy [28]; structural programs that reduce financial strain and food insecurity, including WIC, similarly address allostatic load components that are biologically plausible contributors to LTL attrition and PTB risk [20]. The growing evidence that mindfulness-based interventions are associated with preserved or increased LTL in non-pregnant populations [27,29] provides a biologically plausible rationale for prenatal stress reduction as a strategy that may operate on the same cellular aging pathways indexed by LTL, though this remains to be directly tested in pregnant populations.

### Strengths and Limitations

The principal strengths of this study include its population-based design, confirmed gestational dating by first-trimester ultrasound, the use of the California Biobank enabling LTL measurement in a representative Hispanic/Latina birth sample at a scale not previously available for this question, and access to linked administrative data enabling adjustment for a broad range of demographic and clinical factors. The mid-pregnancy measurement window is a specific strength: by measuring LTL at 15 to 20 weeks, prior to the onset of the clinical complications most strongly associated with PTB, this study captures a pre-morbid biological signal in a way that postpartum studies cannot.

Several important limitations must be acknowledged. First, the absence of direct measures of psychosocial stress, discrimination, or acculturation level precludes direct testing of the allostatic load model that motivates the analysis [9,10,18,19]. Second, several adjustment variables lie on plausible causal pathways between chronic stress and PTB; [45,46] their inclusion may represent overadjustment, and the observed strengthening with full adjustment may reflect negative confounding. Third, participants were drawn from individuals completing routine second-trimester serum screening, which likely enriches for more complete prenatal care utilization relative to the broader California Hispanic/Latina birth population, potentially attenuating observed effect sizes [33,34]. Fourth, gestational age at blood draw within the 15 to 20 week window could not be assessed as a potential source of variation. Fifth, sample sizes precluded examination of early PTB reliably or by PTB subtype including PPROM [59]. Sixth, leukocyte LTL as measured by qPCR reflects average LTL across cell types and does not capture cell-type composition or the distribution of critically short telomeres, which may be more biologically relevant than mean LTL for predicting senescence-related outcomes [63]. Finally, findings are specific to a California Hispanic/Latina population and may not generalize to other geographic or demographic contexts.

### Next Steps

The inconsistency across existing LTL-PTB studies points to several priorities for future investigation. Prospective studies measuring maternal LTL at multiple gestational windows in the same individuals, alongside direct measures of allostatic load, psychosocial stress, discrimination, and acculturation, would allow direct testing of the cumulative burden model and characterization of when during gestation LTL most informatively reflects PTB risk [54,57]. The Phillippe gestational clock model suggests that future studies combining maternal blood LTL with placental or amniotic membrane LTL may yield stronger predictions of parturition timing [57,58,59]. Replication in larger Hispanic/Latina cohorts with sufficient statistical power to examine PTB subtypes, particularly PPROM, is a priority given the LTL-thrombin-PPROM mechanistic evidence [22,23,24,59]. Expanding to other racial and ethnic groups facing disproportionate stress-related PTB burdens would provide important data for evaluating whether the 10th-percentile pattern generalizes across different stress exposure profiles [53]. The development of standardized, clinically deployable LTL measurement methods remains a barrier to translation, and efforts to establish validated clinical cut-points from large, prospectively designed studies represent an important longer-term goal [64].

## Conclusions

PTB rates among Hispanic/Latina individuals in the US have risen substantially over the past decade, likely driven, at least in part, by accumulating structural and psychosocial stressors that are increasingly well-documented but not yet adequately addressed by existing prevention efforts [8,48]. The evidence from this and prior studies supports LTL as a measurable biological marker of this accumulated stress burden, with the present study adding population-based mid-pregnancy data in a Hispanic/Latina cohort specifically [19,20,30,31,32]. We found that LTL at or below the 10th percentile at 15 to 20 weeks of gestation was associated with more than three times the odds of PTB, a finding consistent with the hypothesis that by mid-gestation, those carrying the heaviest allostatic load have experienced sufficient stress-related cellular aging to elevate their biological risk for preterm delivery [48,49,57]. These findings support the value of continued investment in both upstream stress reduction through programs that address the structural conditions driving LTL attrition and PTB in this population, and in prospective research that tests whether LTL measurement can serve as an actionable biological signal in populations where the clinical need is greatest [8,27,28,48].

## Data Availability

All data produced in the present work are contained in the manuscript.

## References

1. March of Dimes. Prematurity Profile: United States. Arlington, VA: March of Dimes; 2024. Available from: https://www.marchofdimes.org/peristats/reports/united-states/prematurity-profile

2. Ely DM and Driscoll AK. Infant Mortality in the United States, 2023: Data From the Period Linked Birth/Infant Death File. Natl Vital Stat Rep. 2025. doi: 10.15620/cdc/174592

3. Siffel C, Hirst AK, Sarda SP, Kuzniewicz MW, Li DK. The clinical burden of extremely preterm birth in a large medical records database in the United States. Early Hum Dev. 2022;171:105613. doi: 10.1016/j.earlhumdev.2022.105613

4. Osterman MJK, Hamilton BE, Martin JA, Driscoll AK, Valenzuela CP. Births: Final Data for 2023. Natl Vital Stat Rep. 2025;74(1). doi: 10.15620/cdc/175204

5. March of Dimes. Preterm birth rate by race/ethnicity: United States, 2022-2024 average. PeriStats [Internet]. 2026. Available from: https://www.marchofdimes.org/peristats

6. Gould JB, Qin C, Chavez G. National trends in preterm infant mortality in the United States by race and socioeconomic status, 1995-2020. JAMA Pediatr. 2023. doi: 10.1001/jamapediatrics.2023.2956

7. Manuck TA. Racial and ethnic differences in preterm birth: a complex, multifactorial problem. Semin Perinatol. 2017;41(8):511–518. doi: 10.1053/j.semperi.2017.08.010

8. Centers for Disease Control and Prevention, National Center for Health Statistics. Natality public-use data, 2016-2024, on CDC WONDER Online Database. Available from: https://wonder.cdc.gov/natality-current.html. Accessed March 5th, 2026.

9. Shapiro GD, Fraser WD, Frasch MG, Seguin JR. Psychosocial stress in pregnancy and preterm birth: associations and mechanisms. J Perinat Med. 2013;41(6):631–645. doi: 10.1515/jpm-2012-0295

10. Wadhwa PD, Entringer S, Buss C, Lu MC. The contribution of maternal stress to preterm birth: issues and considerations. Clin Perinatol. 2011;38(3):351–384. doi: 10.1016/j.clp.2011.06.007

11. Gemmill A, Catalano R, Casey JA, Karasek D, Alcala HE, Elser H, et al. Association of preterm births among US Latina women with the 2016 presidential election. JAMA Netw Open. 2019;2(7):e197084. doi: 10.1001/jamanetworkopen.2019.7084

12. Nichols TR, Mohatt NV, Mohatt DF. Stress, immigration, and mental health: understanding maternal stress in undocumented immigrants during pregnancy. J Immigr Minor Health. 2018;20(3):725–731.

13. Novak NL, Geronimus AT, Martinez-Cardoso AM. Change in birth outcomes among infants born to Latina mothers after a major immigration raid. Int J Epidemiol. 2017;46(3):839–849. doi: 10.1093/ije/dyw346

14. Coussons-Read ME. Effects of prenatal stress on pregnancy and human development: mechanisms and pathways. Obstet Med. 2013;6(2):52–57. doi: 10.1177/1753495X12473751

15. Misra DP, Strobino DM, Trabert B. A life course approach to preconceptional stress and preterm birth. J Perinat Med. 2010;38(2):131–140. doi: 10.1515/jpm.2009.154

16. Epel ES, Blackburn EH, Lin J, Dhabhar FS, Adler NE, Morrow JD, et al. Accelerated telomere shortening in response to life stress. Proc Natl Acad Sci USA. 2004;101(49):17312–17315. doi: 10.1073/pnas.0407162101

17. Barnes RP, Fouquerel E, Opresko PL. The impact of oxidative DNA damage and stress on telomere homeostasis. Mech Ageing Dev. 2019;177:37–45. doi: 10.1016/j.mad.2018.03.009

18. Ruiz RJ, Trzeciakowski J, Moore T, Ayers KS, Pickler RH. Acculturation predicts negative affect and shortened telomere length. Biol Res Nurs. 2017;19(1):28–35. doi: 10.1177/1099800416672005

19. Incollingo Rodriguez AC, Polcari JJ, Nephew BC, Murgatroyd C, Bhatt D, Hale TM, et al. Acculturative stress, telomere length, and postpartum depression in Latinx mothers. J Psychiatr Res. 2022;147:301–306. doi: 10.1016/j.jpsychires.2022.01.063

20. Prasad A, Lin J, Jelliffe-Pawlowski L, Coleman-Phox K, Rand L, Wojcicki JM. Sub-optimal maternal gestational gain is associated with shorter leukocyte telomere length at birth in a predominantly Latinx cohort of newborns. Matern Health Neonatol Perinatol. 2023;9:14. doi: 10.1186/s40748-023-00167-z

21. Wojcicki JM, Sahota M, Lin J, Coleman-Phox K, Jelliffe-Pawlowski L, Rand L. Maternal hypertension and telomere length are associated with weight for age Z score change from birth to 6 months of age in a predominantly Latinx cohort. BMC Pregnancy Childbirth. 2025;25:1195. doi: 10.1186/s12884-025-08213-8

22. Goswami M, Bhagat S, Kumar P, Singhal A. Telomere length and its association with coagulation parameters: a cross-sectional study. Blood Coagul Fibrinolysis. 2023;34(3):181–187. doi: 10.1097/MBC.0000000000001197

23. Hemker HC, Giesen P, Al Dieri R, Regnault V, de Smedt E, Wagenvoord R, et al. Calibrated automated thrombin generation measurement in clotting plasma. Pathophysiol Haemost Thromb. 2007;33(1):4–15.

24. Feng L, Allen TK, Marinello WP, Murtha AP. Role of thrombin in fetal membrane weakening and rupture. J Clin Endocrinol Metab. 2018;103(4):1446–1457. doi: 10.1210/jc.2017-02576

25. Panelli DM, Bianco K. Cellular aging and telomere dynamics in pregnancy. Curr Opin Obstet Gynecol. 2022;34(2):57–61. doi: 10.1097/GCO.0000000000000765

26. Platts S, Bennett PR, Khullar V, Bhide P. Leukocyte telomere length at the first trimester of pregnancy is associated with preterm birth in a small prospective pilot study. BJOG. 2018;125(10):1306–1311. doi: 10.1111/1471-0528.15237

27. Dunn SL, Dimolareva M. The effect of mindfulness-based interventions on immunity-related biomarkers including telomere length and telomerase activity: a systematic review and meta-analysis of randomised controlled trials. Ageing Res Rev. 2022;79:101645. doi: 10.1016/j.arr.2022.101645

28. Janssen LAE, Gieskes AAA, Kok M, de Groot CJM, Oudijk MA, de Boer MA. Stress-reducing interventions in pregnancy for the prevention of preterm birth: a systematic review and meta-analysis. J Psychosom Obstet Gynaecol. 2023;44(1):2281238. doi: 10.1080/0167482X.2023.2281238

29. Mathur MB, Epel E, Kind S, Desai M, Parks CG, Sandler DP, et al. Perceived stress and telomere length: a systematic review, meta-analysis, and methodologic considerations for advancing the field. Brain Behav Immun. 2016;54:158–169. doi: 10.1016/j.bbi.2016.02.002

30. Page RL, Han G, Akinlotan M, Patron MP, Gandhi H, Kochan KJ. Telomere length and preterm birth in pregnant Mexican-origin women. Matern Child Health J. 2021;25(11):1798–1805. doi: 10.1007/s10995-021-03209-0

31. Dutson U, Lin J, Jelliffe-Pawlowski LL, Coleman-Phox K, Rand L, Wojcicki JM. The association between longer maternal leukocyte telomere length in the immediate postpartum period and preterm birth in a predominately Latina cohort of mothers. Matern Child Health J. 2025;29:415–427. doi: 10.1007/s10995-025-04056-z

32. Panelli DM, Wang X, Mayo J, Iqbal SN, Girsen AI, Lyell DJ, et al. Association of pregnancy complications and postpartum maternal leukocyte telomeres in two diverse cohorts: a nested case-control study. BMC Pregnancy Childbirth. 2024;24:490. doi: 10.1186/s12884-024-06688-5

33. Jelliffe-Pawlowski LL, Rand L, Bedell B, Baer RJ, Oltman SP, Norton ME, et al. Prediction of preterm birth with and without preeclampsia using mid-pregnancy immune and growth-related molecular factors and maternal characteristics. J Perinatol. 2018;38(8):963–972. doi: 10.1038/s41372-018-0112-0

34. Smith CJ, Jasper EA, Baer RJ, Breheny PJ, Paynter RA, Bao W, et al. Genetic risk scores for maternal lipid levels and their association with preterm birth. Lipids. 2019;54(10):641–650. doi: 10.1002/lipd.12186

35. Kovac U, Jasper EA, Smith CJ, Baer RJ, Bedell B, Donovan BM, et al. The association of polymorphisms in circadian clock and lipid metabolism genes with second trimester lipid levels and preterm birth. Front Genet. 2019;10:540. doi: 10.3389/fgene.2019.00540

36. Ross KM, Baer RJ, Ryckman K, Feuer SK, Bandoli G, Chambers C, et al. Second trimester inflammatory and metabolic markers in women delivering preterm with and without preeclampsia. J Perinatol. 2019;39(2):314–320. doi: 10.1038/s41372-018-0275-8

37. Bandoli G, Jelliffe-Pawlowski LL, Feuer SK, Liang L, Oltman SP, Paynter R, et al. Second trimester serum cortisol and preterm birth: an analysis by timing and subtype. J Perinatol. 2018;38(8):973–981. doi: 10.1038/s41372-018-0128-5

38. Wallenstein MB, Jelliffe-Pawlowski LL, Yang W, Carmichael SL, Stevenson DK, Ryckman KK, et al. Inflammatory biomarkers and spontaneous preterm birth among obese women. J Matern Fetal Neonatal Med. 2016;29(20):3317–3322. doi: 10.3109/14767058.2015.1124083

39. Monangi NK, Xu H, Fan YM, Khanam R, Khan W, Deb S, et al. Association of maternal prenatal copper concentration with gestational duration and preterm birth: a multicountry meta-analysis. Am J Clin Nutr. 2024;119(1):221–231. doi: 10.1016/j.ajcnut.2023.10.011

40. Monangi N, Xu H, Khanam R, Khan W, Deb S, Pervin J, et al. Association of maternal prenatal selenium concentration and preterm birth: a multicountry meta-analysis. BMJ Glob Health. 2021;6(9):e005856. doi: 10.1136/bmjgh-2021-005856

41. Panelli DM, Li M, Harris LK, Moore JL, Papageorghiou AT, Bhide P, et al. Telomere length: associations with pregnancy complications and placental epigenetics. Reprod Sci. 2022;29(10):2961–2967. doi: 10.1007/s43032-022-00959-6

42. Niu Z, Ding L, Feng Y, Zhang X, Shen Y, Xue M, et al. Differences in leukocyte telomere length, associated with racial/ethnic background, birth characteristics, sex, and age. Eur J Epidemiol. 2019;34(3):255–265. doi: 10.1007/s10654-018-0454-4

43. Burris HH, Hacker MR. Birth outcome racial disparities: a result of intersecting social and environmental factors. Semin Perinatol. 2017;41(6):360–366. doi: 10.1053/j.semperi.2017.07.002

44. Kilpatrick SJ, Papile LA, Macones GA, editors. Guidelines for Perinatal Care. 8th ed. Elk Grove Village, IL: American Academy of Pediatrics; 2016.

45. van Zwieten A, Tennant PWG, Kelly-Irving M, Blyth FM, Teixeira-Pinto A, Khalatbari-Soltani S. Avoiding overadjustment bias in social epidemiology through appropriate covariate selection: a primer. J Clin Epidemiol. 2022;149:127–136. doi: 10.1016/j.jclinepi.2022.05.021

46. Lu H, Cole SR, Platt RW, Schisterman EF. Revisiting overadjustment. Epidemiology. 2021;32(5):e22-e23. doi: 10.1097/EDE.0000000000001377

47. Cawthon RM. Telomere measurement by quantitative PCR. Nucleic Acids Res. 2002;30(10):e47. doi: 10.1093/nar/30.10.e47

48. Premji SS, Pana GS, Cuncannon A, Ronksley PE, Dosani A, Hayden KA, et al. Prenatal allostatic load and preterm birth: a systematic review. Front Psychol. 2022;13:1004073. doi: 10.3389/fpsyg.2022.1004073

49. Rentscher KE, Carroll JE, Mitchell C. Psychosocial stressors and telomere length: a current review of the science. Annu Rev Public Health. 2020;41:223–245. doi: 10.1146/annurev-publhealth-040119-094239

50. Geronimus AT, Hicken M, Keene D, Bound J. Weathering and age patterns of allostatic load scores among Blacks and Whites in the United States. Am J Public Health. 2006;96(5):826–833. doi: 10.2105/AJPH.2004.060749

51. Collins JW Jr, Rankin KM, Hedstrom AB. Lifetime upward economic mobility and US-born Latina women’s preterm birth rates. Matern Child Health J. 2024. doi: 10.1007/s10995-023-03890-3

52. Alhusen JL, Bower KM, Epstein E, Sharps P. Racial discrimination and adverse birth outcomes: an integrative review. J Midwifery Womens Health. 2016;61(6):707–720. doi: 10.1111/jmwh.12490

53. Huang W, Han G, Taylor BD, Neal G, Kochan K, Page RL. Maternal peripheral blood telomere length and preterm birth in African American women: a pilot study. Arch Gynecol Obstet. 2025;311:1591–1598. doi: 10.1007/s00404-024-07681-1

54. Schneper LM, Brooks-Gunn J, Notterman DA, Suomi SJ. Leukocyte telomere dynamics across gestation in uncomplicated pregnancies and associations with stress. BMC Pregnancy Childbirth. 2022;22:483. doi: 10.1186/s12884-022-04693-0

55. Cherkas LF, Aviv A, Valdes AM, Hunkin JL, Gardner JP, Surdulescu GL, et al. The effects of social status on biological aging as measured by white-blood-cell telomere length. Aging Cell. 2006;5(5):361–365. doi: 10.1111/j.1474-9726.2006.00222.x

56. Needham BL, Adler N, Gregorich S, Rehkopf D, Lin J, Blackburn EH, et al. Socioeconomic status, health behavior, and leukocyte telomere length in NHANES, 1999-2002. Soc Sci Med. 2013;85:1-8. doi: 10.1016/j.socscimed.2013.02.023

57. Phillippe M. Telomeres, oxidative stress, and timing for spontaneous term and preterm labor. Am J Obstet Gynecol. 2022;227(2):148–162. doi: 10.1016/j.ajog.2022.04.024

58. Polettini J, Dutta EH, Behnia F, Saade GR, Torloni MR, Menon R. Telomere-related disorders in fetal membranes associated with birth and adverse pregnancy outcomes. Front Physiol. 2020;11:561771. doi: 10.3389/fphys.2020.561771

59. Menon R, Yu J, Basanta-Henry P, Brou L, Bhatt DL, Fortunato SJ, et al. Short fetal leukocyte telomere length and preterm prelabor rupture of the membranes. PLoS One. 2012;7(2):e31136. doi: 10.1371/journal.pone.0031136

60. American College of Obstetricians and Gynecologists. Preterm labor and birth. ACOG Practice Bulletin No. 234. Obstet Gynecol. 2021;138(6):e65-e90. doi: 10.1097/AOG.0000000000004658

61. Fryer K, Lewis G, Munoz C, Stuebe AM. Identifying barriers and facilitators to prenatal care for Spanish-speaking women. N C Med J. 2021;82(1):7–13. doi: 10.18043/ncm.82.1.7

62. Ortiz K, Garcia A, Salazar C, Saulsberry L, Beal SJ, Wingrove PM, et al. Perinatal care among Hispanic birthing people: differences by primary language and state policy environment. Birth. 2024. doi: 10.1111/birt.12830

63. Ahrens KA, Rossen LM, Simon AE. Relationship between mean leucocyte telomere length and measures of allostatic load in US reproductive-aged women, NHANES 1999-2002. Paediatr Perinat Epidemiol. 2016;30(4):325-335. doi: 10.1111/ppe.12277

64. Blackburn EH, Epel ES, Lin J. Human telomere biology: a contributory and interactive factor in aging, disease risks, and protection. Science. 2015;350(6265):1193–1198. doi: 10.1126/science.aab3389

